# Association of Abnormal Cytology and risk factors in Women of Assam, India

**DOI:** 10.1101/2023.01.16.23284644

**Authors:** Debabrata Barmon, Anupam Sarma, Upasana Baruah, Lopamudra Kakoti, Debanjana Barman, Pallavi Sarma, Manoj Kalita, Avdhesh Kumar Rai

## Abstract

The study was conducted to perform cytological abnormality screening by PAP test among women of Assam, India. Two hundred thirty four women in the age group of 35-60 years were enrolled in screening camps for the study. Smear on the glass slide of the cervical scrap specimen were prepared for PAP test. Statistical analysis for PAP test outcome with risk factors was analyzed by applying Fisher’s exact test. Significance was considered for p<0.05.

It was found that women using contraceptive piles have higher risk of abnormal cytology (p-0.0238). Women with irregular menstrual cycle frequency have also higher risk of abnormal cytology (p-0.0002). Women not using commercially available sanitary pads ahe also high risk of abnormal cytology( p-0.0052). Women having urinary tract infection have also high risk of abnormal cytology (p-0.0102). Our study finding suggest that use of contraceptive pills, urinary infection, irregular frequency of menstrual cycle, non-use of sanitary pads may increase the risk of abnormal cytology positivity in women of Assam, India. There are limitations of our studyfindings because of small sample size and univariate statistical analysis.

## INTRODUCTION

Cervical cancer has remained one of the worst mortality factors among women suffering from cancer globally and especially in India. About a quarter of incidence and mortality due to cervical cancer of global level was reported from India in 2020^1^. Cervical cancer is one of the most preventable cancer if complemented with appropriate screening by Papanicolaou tests (PAP) test will lead to early detection of precancerous cervical lesions. It has been well documented from evidence based studies across globe that PAP test effective implementation has been successful in drastically reducing incidence and mortality due to invasive cervical cancer ^2^.

PAP cytology test has been found to be most easy, cost effective, implementable principal screening technique for identification of cervical Intraepithelial neoplasia lesions(CINs) in industrialized countries (Europe and USA) who implemented it as part of their national CINs screening policy^3^. Developing countries under the category of Low and middle income countries are using PAP test as primary screening method for CINs but still they are facing hurdles to adopt as mandatory national screening programme among women^4^.

Cervical cancer screening programmes need to develop starategy which is tandardized for developing countries participating women with taken in consideration socio-cultural dynamics of the societies of the participating women. This will be help better follow up outcome for PAP test and subsequent mitigation plan to reduce incidence of invasive cervical cancer in screened women population ^5^.

Cervical cancer age adjusted incidence rate (AAR) per 100000 population of north-east India was reported to be as high as 30 ^6^. This study was conducted in one district of Assam, India to assess the prevalence of abnormal cytology by PAP test among women for CINs and PAP test outcome association with risk factors.

## MATERIALS AND METHOD

Screening cum awareness camps for women about cervical cancer prevention were orgazined in a district of Assam, India. Participating women who were eligible for screening were in the age group of 30-65 years. Women were informed about the consenting process and provided informed written consent. Total eligible women participant number was two hundred thirty four (n=234). In a detailed proforma questionnaire, information about risk factors from women was collected by process of personnel interaction. The Institutional Ethics Committee has approved this study.

### Papanicolaou (PAP) Test

Cervical scrap specimen collected from the cervical region of the women with soft cervical cytological purpose brush. The brush was rotated five times in clock and counter clockwise direction inside the women cervix. The brush was then rolled over thoroughly on the clean surface of glass slide in smear preparation manner. The cervical cell scrap smear was fixed with 95% ethanol for 15 min. Slides were gently rinsed in deionized water for 30 sec. Staining in hematoxylin solution was doen for 2 min. After staining, slides were again rinsed in deionized water for 1 min. Slides were incubated in bluing reagent for 1 min. slides were again rinsed in deionized water. Slides were dipped for 5-10 times in 95% ethanol. Slides were placed in Papanicolaou Stain EA-50/ EA-65 solution for 2-3min. Slides were fixed in 95% ethanol. Slides were visualized under bright filed microscope for cytological abnormality assessment and reporting. For assessment and reporting of PAP test outcome the Bethesda system for reporting for cytology was used. Positive PAP test scoring and reporting was done all the samples which have following morphological features: HSIL, LSIL and ASCUS.

### Statistical Analysis

Abnormal cytology outcome association with risk factor done using Fisher exact test. Statistivcal analysis was performed in Graphpad prism software version10. Statistical significance was considered at p<0.05.

## RESULTS

Among the screened women, 5.12% women 912/234) PAP smears had shown abnormal cytology morphological features for ASCUS, LSIL, HSIL. Association between abnormal cytology with risk factors is elaborated. Women who used contraceptive piles have shown significant association with positive abnormal cytology (p-0.0238). Women with irregular frequency of menstrual cycle has also shown significant risk for positive PAP outcome (p-0.0002). Women not using commercial sanitary pads have also shown significantly high risk for positive abnormal cytology (p-0.0052). Women having urinary tract infection have also significantly higher risk of abnormal cytology outcome (p-0.0102). (Table1)

**Table 1:** Abnormal Cytology outcome (PAP Test) with behavioral characteristics of participant women.

| Variables |  | PAP |  | P- Value |
| --- | --- | --- | --- | --- |
|  |  | Negative | positive |  |
| Tobacco Chew | YES | 77 | 7 | 0.1236 |
|  | NO | 145 | 5 |  |
| Betel nut Chew | YES | 148 | 8 | 1.0000 |
|  | NO | 74 | 4 |  |
| Alcohol Consumption | YES | 47 | 4 | 0.3089 |
|  | NO | 174 | 8 |  |
| Use of contraceptives pills | YES | 71 | 8 | 0.0238* |
|  | NO | 151 | 4 |  |
| Menstrual Cycle Frequency | Regular | 174 | 3 | 0.0002* |
|  | Irregular | 48 | 9 |  |
| Use of Commercial Sanitary Pad | YES | 178 | 5 | 0.0052* |
|  | NO | 44 | 7 |  |
| Urinary tract infection of spouse | YES | 88 | 6 | 0.5510 |
|  | NO | 134 | 6 |  |
| Urinary Tract Infection of Self | YES | 77 | 9 | 0.0102* |
|  | NO | 145 | 3 |  |

\*Statistically Significant

## DISCUSSION

PAP positivity in our study was found to be 5.12 %. The range for PAP positivity was reported to be 0.6-4.5% reflecting wide variations within and across geological regions ^7,8^. PAP smear cytology have certain limitation with regard to individual observation bias which may be related to experience in cytology based methodology.

We found in our study that Women using contraceptives have shown significantly higher risk for positive abnormal cytology (p-0.0238). It was reported that long term use(more than 10 years of oral contraceptive pills have increased the risk of cervical cancer in women^9^. A large study from Japan found that women who used injectable contraceptive for five years, have manifold higher risk of cervical cancer^10^. Another study has also suggested that Oral Contraceptive pills may be significant predictor for high-grade CINs lesions^11^. Meta analysis of more than two dozen studies has concluded that women who used oral contraceptive pills have increased risk of cervical cancer in their life time ^12^. A large study has not found any significant association between oral contraceptive pills use and abnormal PAP outcome ^13^. Inconsistency between the association of oral contraceptive pills use and abnormal PAP outcome among various studiesmay be due to multiple confounding life style factors.

We had also found in our study that women who did not use commercial sanitary pad has shown significantly higher risk for positive abnormal cytology outcome (p-0.0052). Sri Syatriani’s (2011) reported that use of low quality sanitary pads can significantly increase the risk of abnormal cytology and cervical cancer in women^14^. It was also suggested that clothes reused as sanitary pads increase risk of positive abnormal cytology outcome and cervical cancer^12^. We found that women having irregular frequency of menstrual cycle (p-0.0002) and urinary tract infection (0.0102) has significantly higher risk of positive abnormal cytology outcome. It was reported that poor genital hygiene and abnormal menstruation were found to be significantly associated with increased risk of CINs ^15,16^.

Women of Assam need to made more aware about good genital hygiene practice( use of good quality sanitary napkins, safe sex practices to protect against urinary tract infection and factors associated with reproductive health such as regularity of menstrual cycle to reduce the risk for CINs.

## CONCLUSION

On the basis of our current findings we conclude that use of oral contraceptive pills, irregular frequency of menstrual cycle, non use of good quality sanitary pads, urinary tract infection significantly associated with positive abnormal cytology outcome by PAP test. Future studies with consideration for other risk factors such as HPV with multi centre study design will be required.

## Data Availability

All data is available in manuscript

## ACKNOWLEDGEMENT

Authors express their gratitude

## CONFLICT OF INTEREST STATEMENT

All authors declare that they have no affiliations with or involvement in any organization or entity with any financial interest or non-financial interest in the subject matter or materials discussed in this manuscript.

## Notes

### Competing Interest Statement

The authors have declared no competing interest.

### Funding Statement

This study didn't receive funding

### Author Declarations

Ethics committee/IRB of Dr Bhubaneswar Borooah Cancer Institute, Guwahati, Assam, INDIA gave ethical approval for this work

### Summary of Updates

This manuscript has been revised to update statistical outcome and the manuscript introduction, methods, results, discussion, conclusion, reference, abstract and author list also has been revised.

## REFERENCES

1. Sung H, Ferlay J, Siegel R L, Laversanne M et al. Global cancer statistics 2020: GLOBOCAN estimates of incidence and mortality worldwide for 36 cancers in 185 countries. Ca Cancer J Clin 2021; 71:209–249.

2. Zhang, J., Zhao, Y., Dai, Y., Dang, L., Ma, L., Yang, C. et al. Effectiveness of High-risk Human Papillomavirus Testing for Cervical Cancer Screening in China: A Multicenter, Open-label Randomized Clinical Trial. JAMA oncology 2021 ;7(2) : 263–270.

3. Jemal A, Center MM, DeSantis C, Ward EM. Global patterns of cancer incidence and mortality rates and trends. Cancer Epidemiol Biomarkers Prev. 2010;19(8):1893–907.

4. Dykens J A, Smith J S, Demment M et al. Evaluating the implementation of cervical cancer screening programs in low-resource settings globally: a systematized review. Cancer Causes Control 2020; 31: 417–429.

5. Arbyn M, Smith SB, Temin S et al. Detecting cervical precancer and reaching underscreened women by using HPV testing on self samples: updated meta-analyses. BMJ 2018; 363:k4823.

6. Indian Council of Medical Research, National Cancer Registry programme: Three Year Report of Population based Cancer Registries 2012-2014 (Report of 27 PBCRs in India). 2016 https://www.ncdirindia.org/All_Reports/PBCR_REPORT_2012_2014

7. Dey S, Pahwa P, Arti Mishra et al. Reproductive Tract infections and Premalignant Lesions of Cervix: Evidence from Women Presenting at the Cancer Detection Centre of the Indian Cancer Society, Delhi, 2000-2012. J ObstetGynaecolIndia 2016 ; 66(Suppl 1):441–51.

8. Mulay K, Swain M, Patra S et al. A comparative study of cervical smears in an urban Hospital in India and a population-based screening program in Mauritius. Indian J Pathol Microbiol 2009 ; 52:34–37.

9. Brinton LA. Oral contraceptives and cervical neoplasia. Contraception. 1991;43:581–595.

10. Herrero R, Brinton LA, Reeves WC, Brenes MM, De Britton RC, Tenorio F, et al. Injectable contraceptives and risk of invasive cervical cancer: evidence of an association. Int J Cancer. 2006;46:5–7.

11. Kiatiyosnusorn R, Suprasert P, Srisomboon J, Siriaree S, Khunamornpong S, Kietpeerakool C. High-grade histologic lesions in women with low-grade squamous intraepithelial lesion cytology from a region of Thailand with a high incidence of cervical cancer. Int J Gynaecol Obstet. 2010;110:133–136.

12. P. Appleby, V. Beral, B. A. de Gonzalez, and D. Colin. Cervical cancer prevention and hormonal contraception . Lancet 2007 ; vol. 370, no. 9599, pp. 1591–1592.

13. Binesh F, Akhavan A, Pirdehghan A, Davoodi M. Does oral contraceptive pill increase the risk of abnormal Pap smear? Iran J Reprod Med. 2013 Sep;11(9):761–6.

14. SYATRIANI, Sri. Faktor Risiko Kanker Serviks di Rumah Sakit Umum Pemerintah Dr. Wahidin Sudirohusodo Makassar, Sulawesi Selatan. Kesmas: Jurnal Kesehatan Masyarakat Nasional (National Public Health Journal), [S.l.], p. 283–288, june 2011.

15. Juneja A, Sehgal A, Mitra AB, Pandey A (2003). A survey on risk factors with cervical cancer. Indian J Cancer, 40, 15–22.

16. Reiter PL, Katz ML, Ferketich AK, et al (2009). Measuring cervical cancer risk:developmet and validation of the care risky sexual behavior index. Cancer Causes and Control, 20, 1865–71.

